# Temporal Associations between Community Incidence of COVID-19 and Nursing Home Outbreaks in Ontario, Canada

**DOI:** 10.1101/2020.11.17.20233312

**Authors:** Kamil Malikov, Qing Huang, Shengli Shi, Nathan M. Stall, Ashleigh R. Tuite, Michael P. Hillmer

**Affiliations:** Capacity Planning and Analytics Division, Ontario Ministry of Health, Toronto, Canada; Department of Medicine, University of Toronto, Toronto, Canada; Institute of Health Policy, Management and Evaluation, University of Toronto, Toronto, Canada; Women’s College Research Institute, Women’s College Hospital, Toronto, Canada; Division of General Internal Medicine and Geriatrics, Sinai Health System and the University Health Network, Toronto, Canada; Dalla Lana School of Public Health, University of Toronto, Toronto, Canada

**Author notes:** **Corresponding author:** Michael P. Hillmer MSc, PhD, Assistant Deputy Minister, Capacity Planning and Analytics Division, Ontario Ministries of Health and Long-Term Care, 1075 Bay Street, 13th Floor, Toronto, Ontario M5S 2B1 Canada.

## Abstract

The risk of nursing home COVID-19 outbreaks is strongly associated with the rate of infection in the communities surrounding homes, yet the temporal relationship between rising rates of community COVID-19 infection and the risk threshold for subsequent nursing home COVID-19 outbreaks is not well defined. This population-based cohort study included all COVID-19 cases in Canada’s most populous Province of Ontario between March 1-July 16, 2020. We evaluated the temporal relationship between trends in the number of active community COVID-19 cases and the number of nursing home outbreaks. We found that the average lag time between community cases and nursing home outbreaks was 23 days for Ontario overall, with substantial variability across geographic regions. We also determined thresholds of community incidence of COVID-19 associated with a 75% probability of observing a nursing home outbreak 5, 10 and 15 days into the future. For the province overall, when daily active COVID-19 community cases are 2.30 per 100,000 population, there is a 75% probability of a nursing home outbreak occurring five days later.

## Background

Nursing homes have borne the brunt of the COVID-19 pandemic, with residents of these homes incurring extreme morbidity and mortality (1). The risk of nursing home COVID-19 outbreaks is strongly associated with the rate of infection in the communities surrounding homes, with infected health care workers being important and unknowing vectors for transmission into homes (2, 3). The temporal relationship between rising rates of community COVID-19 infection and the risk threshold for subsequent nursing home COVID-19 outbreaks is not well defined.

### Objective

Evaluate and quantify the temporal relationship between community incidence of COVID-19 and subsequent risk of COVID-19 outbreaks in nursing homes in Ontario, Canada.

## Methods and Findings

This population□based cohort study included all laboratory-confirmed COVID-19 cases in the Province of Ontario, Canada (population >14-million) between March 1, 2020 (start of community transmission of COVID-19) and July 16, 2020 (no new nursing home outbreak for >7-day period). We obtained data for this study from the Ontario Ministry of Health as part of the province’s emergency “modeling table,” including deidentified line level data from the integrated Public Health Information System on all reported COVID□19 cases for both community and nursing home dwelling Ontario residents. We also obtained data on COVID-19 outbreaks from the province’s Long-Term Care Inspections Branch COVID-19 case tracking tool. In Ontario, a nursing home COVID-19 outbreak is defined as either one resident or staff case and is declared over when there are no news cases within a 14-day period. All statistical analyses were completed in SAS Statistical Software and Python. The study was approved by the Research Ethics Board of the University of Toronto.

There was a total of 37,274 COVID-19 cases reported over this time period, of which 5,545 (14.8%) were reported among residents of 343 cumulative nursing home outbreaks. We assigned all nursing homes to one of Ontario’s five administrative health regions (West, Central, East, North, and Toronto). We then evaluated the temporal relationship between trends in the number of active community COVID-19 cases (cumulative cases less resolved cases and deaths) in each geographic region and the number of nursing home outbreaks (**Figure 1**). Active cases were used because they are better reflective of the risk of infection in the population. We calculated Pearson’s correlation coefficients (r) between the number of nursing home outbreaks and daily active community cases of COVID-19 in the days (1-50) preceding outbreaks, and ranked coefficients by their descending values. The day with highest r value was chosen as the ‘lag day’ indicator. The average lag time between community cases and nursing home outbreaks was 23 days for Ontario overall, with substantial variability across geographic regions ranging from 11 to 43 days (**Table 1**). The longest lag was observed in the North Region, which has low population density and reported a substantially lower cumulative COVID-19 incidence in nursing home residents (0.3%) compared to the provincial average (7.5%) over this time period.

**Table 1:** Characteristics of Ontario’s 623 nursing homes and lag times between community incidence of COVID-19 and nursing home outbreaks (March 1-July 16, 2020)

| Geographic region | Population density per square kilometre<sup>a</sup> | Number of nursing homes | Number of nursing home residents | Cumulative COVID-19 cases in residents | Threshold of daily active COVID-19 community cases per 100,000 population<sup>b</sup> resulting in a 75% probability of a future nursing home outbreak |  |  |
| --- | --- | --- | --- | --- | --- | --- | --- |
|  |  |  |  |  | 5 days | 10 days | 15 days |
| Central | 294.4 | 123 | 17,315 | 2,273 | 2.93 | 4.02 | 5.84 |
| East | 64.7 | 165 | 20,327 | 1,936 | 1.88 | 5.53 | 6.38 |
| North<sup>c</sup> | 1.0 | 63 | 6,495 | 17 | – | – | – |
| Toronto | 6,412.6 | 36 | 5,695 | 739 | 3.52 | 9.63 | 13.58 |
| West | 94.9 | 236 | 25,844 | 716 | 1.83 | 3.07 | 1.64 |
| Ontario | 14.8 | 623 | 75,676 | 5,681 | 2.30 | 3.65 | 3.93 |

<sup>a</sup> Source: Ontario Ministry of Finance, based on 2016 Canadian Census.
<sup>b</sup> Daily active COVID-19 community cases per 100,000 population. All active COVID-19 community cases (cumulative cases less resolved cases and deaths) and COVID-19 nursing home outbreaks in the Province of Ontario were calculated during the period from March 1 – April 8, 2020, after which we conducted logistic regression on all active cases and nursing home outbreaks from April 8 – July 16, 2020.
<sup>c</sup> Threshold calculations for the North Region were suppressed due to the small numbers of reported cases and small population size.

**Figure 1:**
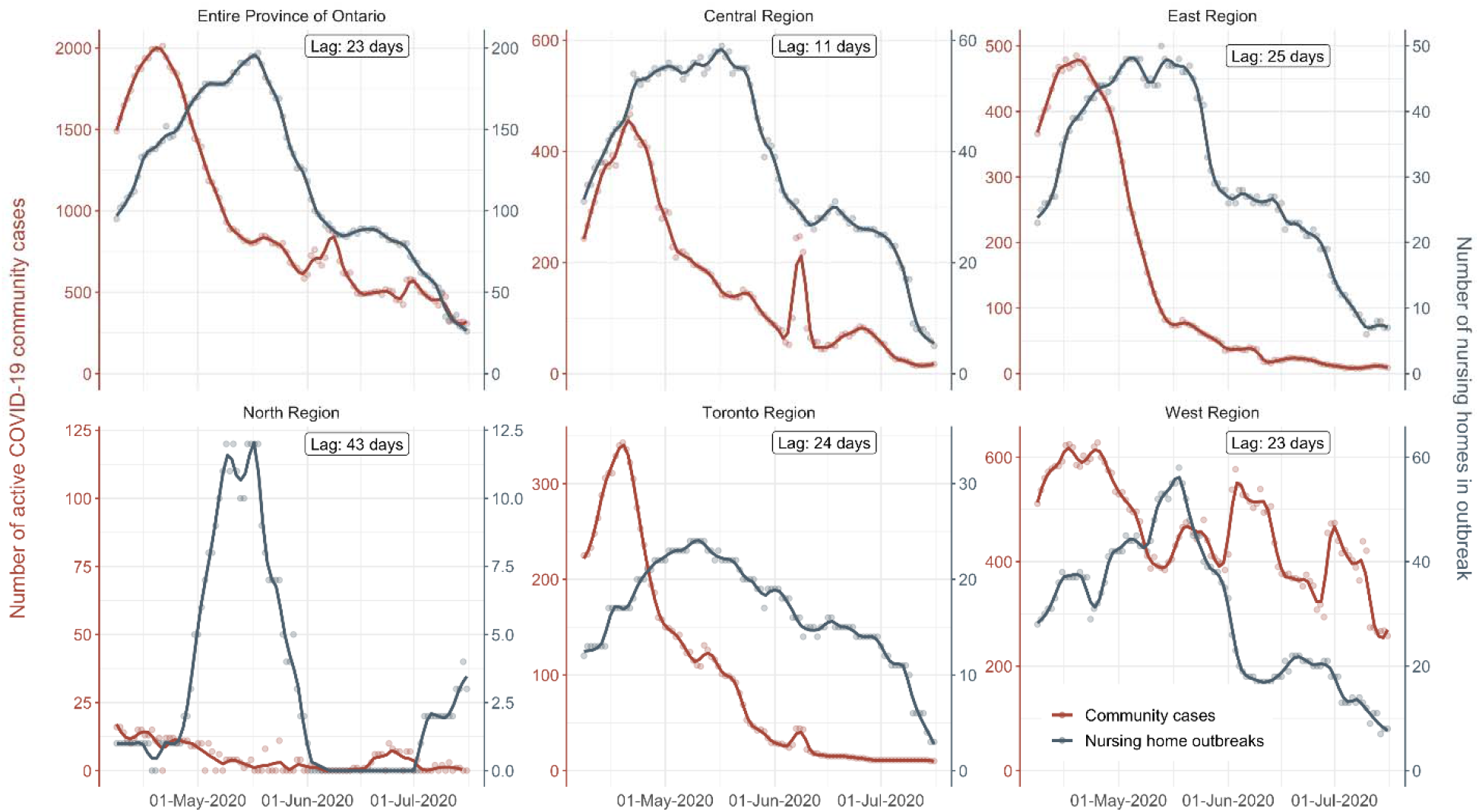
Temporal relationship between community incidence of COVID-19 and outbreaks in Ontario’s 623 nursing homes (April 8-July 16, 2020) All active COVID-19 community cases (cumulative cases less resolved cases and deaths) and COVID-19 nursing home outbreaks in the Province of Ontario were calculated during the period from March 1 – April 8, 2020, after which we evaluated the temporal relationship between cases and outbreaks from April 8 – July 16, 2020. Results are displayed for the entire Province of Ontario and its 5 provincial health regions. The reported lag times indicate the number of days most correlated between active COVID-19 community cases and nursing home outbreaks.

We next used logistic regression to model the probability of a nursing home outbreak with the independent variable being active community COVID-19 cases, as above, in the preceding days before an outbreak. We determined thresholds of community incidence of COVID-19 associated with a 75% probability of observing a nursing home outbreak 5, 10 and 15 days into the future (**Table 1**). For the province overall, when daily active COVID-19 community cases are 2.30 per 100,000 population, there is a 75% probability of a nursing home outbreak occurring five days later.

## Discussion

Across Canada’s most populous province of Ontario, increased community COVID-19 transmission portended a 23-day lagged rise in the number of nursing homes experiencing COVID-19 outbreaks. Our findings also establish thresholds for community infections at which outbreaks in nursing homes first occur. This is a useful early warning indicator when establishing surveillance systems, and the lag days estimate provides a time window during which nursing homes should rapidly mobilize occupational health and infection prevention and control processes to both prevent and mitigate COVID-19 outbreaks.

Our analytic approach reinforces the importance of disaggregating community and nursing home populations in models of COVID-19, and may also be applicable to other congregate care settings including assisted living facilities (4). Our findings are also highly relevant to jurisdictions like the United States who are implementing phased approaches to reopening nursing homes based on COVID-19 case status in the community (5).

## Data Availability

Study protocol and statistical code: The study protocol and underlying analytic code are available from the authors on request (e-mail), with the understanding that the computer programs may rely on coding templates or macros that are unique to the Ontario Ministry of Health and therefore either are inaccessible or may require modification. Data set: The data used for this study are not publicly available.

## References

1. Ouslander JG, Grabowski DC. COVID-19 in Nursing Homes: Calming the Perfect Storm. J Am Geriatr Soc. 2020.

2. Stall NM, Jones A, Brown KA, Rochon PA, Costa AP. For-profit long-term care homes and the risk of COVID-19 outbreaks and resident deaths. CMAJ. 2020;192(33):E946–E55.

3. Fisman DN, Bogoch I, Lapointe-Shaw L, McCready J, Tuite AR. Risk Factors Associated With Mortality Among Residents With Coronavirus Disease 2019 (COVID-19) in Long-term Care Facilities in Ontario, Canada. JAMA Netw Open. 2020;3(7):e2015957.

4. Pillemer K, Subramanian L, Hupert N. The Importance of Long-term Care Populations in Models of COVID-19. JAMA. 2020;324(1):25–6.

5. Centers for Medicare & Medicaid Services 2020;Pageshttps://www.cms.gov/files/document/qso-20-30-nh.pdf-0 on May 18 2020.

